# Inherited rare, deleterious variants in *ATM* increase lung adenocarcinoma risk

**DOI:** 10.1101/2020.03.19.20034942

**Authors:** Myvizhi Esai Selvan, Marjorie G. Zauderer, Charles M. Rudin, Siân Jones, Semanti Mukherjee, Kenneth Offit, Kenan Onel, Gad Rennert, Victor E. Velculescu, Steven M. Lipkin, Robert J. Klein, Zeynep H. Gümüş

## Abstract

**Introduction:** Lung cancer is the leading cause of cancer deaths in the world, and adenocarcinoma (LUAD) is its most prevalent subtype. Symptoms often appear in advanced disease when treatment options are limited. Identifying genetic risk factors will enable better identification of high-risk individuals.

**Methods:** To identify LUAD risk genes, we performed a case-control association study for gene-level burden of rare, deleterious variants (RDVs) in germline whole-exome sequencing (WES) data of 1,083 LUAD patients and 7,650 controls, split into discovery and validation cohorts. Of these, we performed WES on 97 patients and acquired the rest from multiple public databases. We annotated all rare variants for pathogenicity conservatively, using ACMG guidelines and ClinVar curation, and investigated gene-level RDV burden using penalized logistic regression. All statistical tests were two-sided.

**Results:** We discovered and replicated the finding that the burden of germline *ATM* RDVs was significantly higher in LUAD patients versus controls (OR_combined_=4.6; *p*=1.7e-04; 95% CI=2.2–9.5; 1.21% of cases; 0.24% of controls). Germline *ATM* RDVs were also enriched in an independent clinical cohort of 1,594 patients from the MSK-IMPACT study (0.63%). Additionally, we observed that an Ashkenazi Jewish (AJ) founder *ATM* variant, rs56009889, was statistically significantly more frequent in AJ cases versus AJ controls in our cohort (OR_combined, AJ_=2.7, *p*=6.9e-03, 95% CI=1.3–5.3).

**Conclusions:** Our results indicate that *ATM* is a moderate-penetrance LUAD risk gene, and that LUAD may be part of the *ATM*-related cancer syndrome spectrum. Individuals with *ATM* RDVs are at elevated LUAD risk and can benefit from increased surveillance (particularly CT scanning), early detection and chemoprevention programs, improving prognosis.

## INTRODUCTION

Lung cancer is the leading cause of cancer deaths in the USA^1^ and worldwide. Although prognosis is substantially better in early-stage as opposed to late-stage disease, most patients are diagnosed at advanced stages, when treatment options are limited.^2^ Understanding genetic risk factors will help identify high-risk individuals, who can then significantly benefit from intensive surveillance (particularly CT scanning),^3,4^ early detection and precision prevention strategies.^5^

Although smoking is a primary risk factor for lung cancer, only ∼15% of smokers develop the disease.^6^ Non-small cell lung cancer (NSCLC) accounts for approximately 90% of all cases, and lung adenocarcinoma (LUAD) is the most prevalent subtype (40%).^7^ Consistent with a genetic predisposition, some NSCLC patients have a positive family history, and are affected at a young age, although they have never smoked. In fact, a family history of lung cancer increases risk,^8^ and heritability is estimated at 18%.^9^ However, LUAD is not known to be a part of any cancer predisposition syndrome. Furthermore, genome-wide scans for common polymorphisms^10^ have only explained a small fraction of overall heritability, and thus cannot distinguish high-risk individuals. As rare variants are expected to have larger effect size, as we observed for lung squamous cell carcinoma,^11^ we reasoned that a similar approach focused on germline rare deleterious variants (RDVs) will provide novel insights into LUAD risk.

## MATERIALS AND METHODS

### Study design

The study design is summarized in Figure 1 and described in detail below.

**Figure 1:**
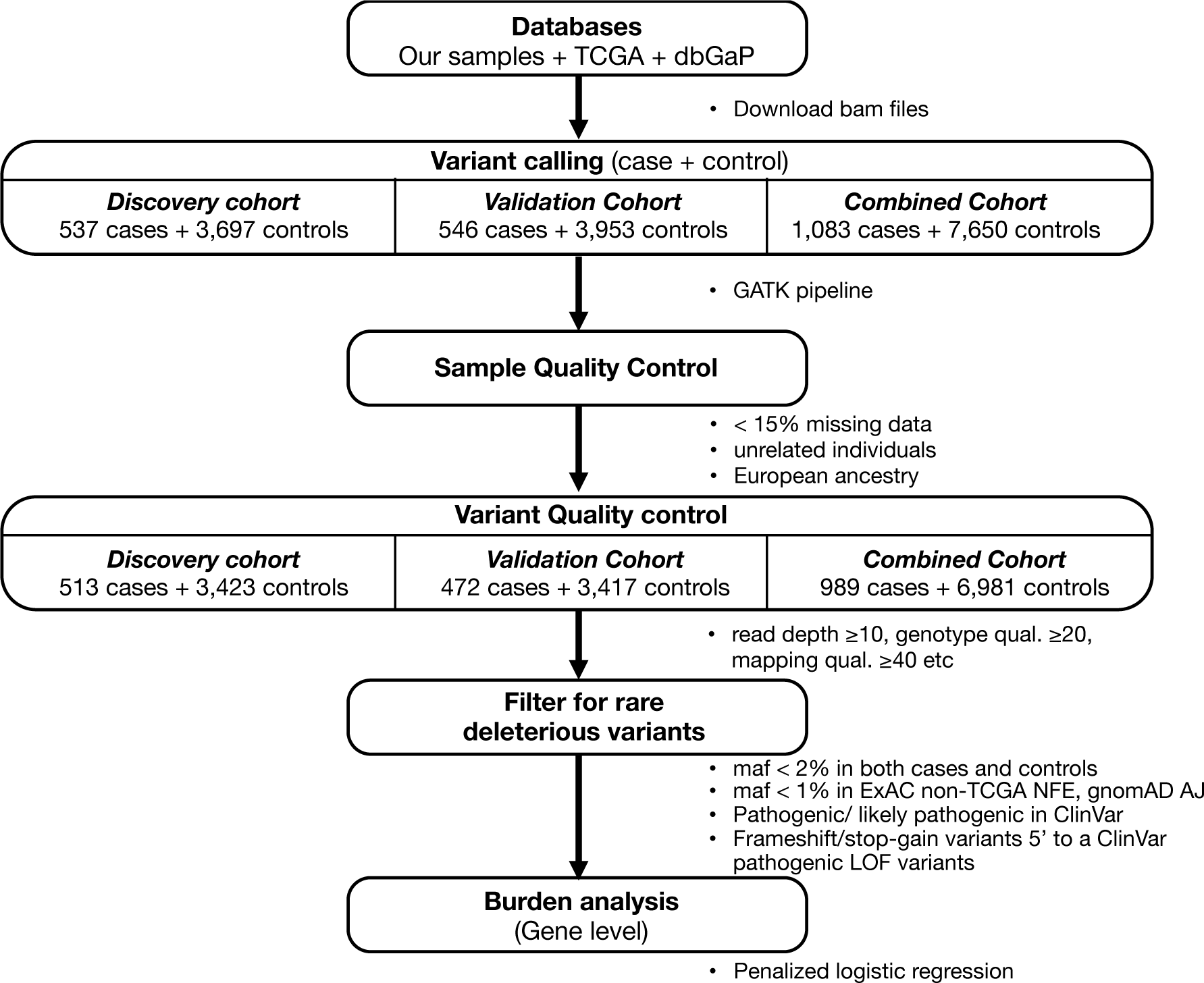
Study design to perform burden analysis.

### Sample collection

We first recruited patients from three NYC institutes in the USA, including Memorial-Sloan Kettering Cancer Center (MSKCC; n=7, IRB #15-061), Weill Cornell Medical College (WCMC; n=14, IRB #1008011221) and Icahn School of Medicine at Mount Sinai (ISMMS; n=2, IRB #12-1072) and from the Lung Cancer in Northern Israel (LCINIS) study conducted at Carmel Medical Center and Clalit National Cancer Control Center (NICCC) in Israel (n=74). This patient cohort was enriched in individuals with familial lung cancer. We collected and processed 97 blood (WCMC; LCINIS) or spit (MSKCC; ISMMS) samples for whole-exome sequencing (WES) under IRB-approved protocols. Sample preparation and WES details are provided in Supplemental Method 1.

### Data acquisition

Pursuing a resource-conscious approach, we analyzed these WES data jointly with already existing germline LUAD WES datasets. Specifically, we added case-control WES data from the Transdisciplinary Research into Cancer of the Lung (TRICL) project, which we downloaded from the database of Genotypes and Phenotypes (dbGaP, www.ncbi.nlm.nih.gov/gap) (phs000876). We then used controls from three population-based studies in dbGaP (ClinSeq project (phs000971); Myocardial Infarction Genetics (MIGen) Exome Sequencing Consortium, U. of Leicester study (phs001000); and Malmo Diet and Cancer Study (phs001101)). We designated this case-control cohort as our *discovery cohort*. For *validation cohort*, we used cases from The Cancer Genome Atlas (TCGA) and controls from eight population-based studies in dbGaP listed in Supplemental Method 2. We downloaded the TCGA germline WES BAM files from National Cancer Institute Genomic Data Commons (GDC) data portal (https://gdc-portal.nci.nih.gov), and control samples from dbGaP.

### Study Cohorts

Despite having WES data for all individuals, to protect against false positives and to ensure reproducibility, we used our WES cases together with TRICL cases for discovery and the TCGA cases for replication cohorts with separate controls, and thereby divided our study into a discovery and a replication cohort (Supplemental Table 1). Overall, for discovery cohort, we analyzed 537 cases (97 we sequenced and 440 from TRICL study) and 3,697 controls (853 from TRICL and 2,844 from ClinSeq and MIGen studies). For validation cohort, we utilized 546 sporadic cases in TCGA and 3,953 controls from eight population-based studies in dbGaP. Together, the combined cohort included 1,083 cases and 7,650 controls. The clinical characteristics of all cohorts after sample QC are listed in Table 1.

**Table 1:**
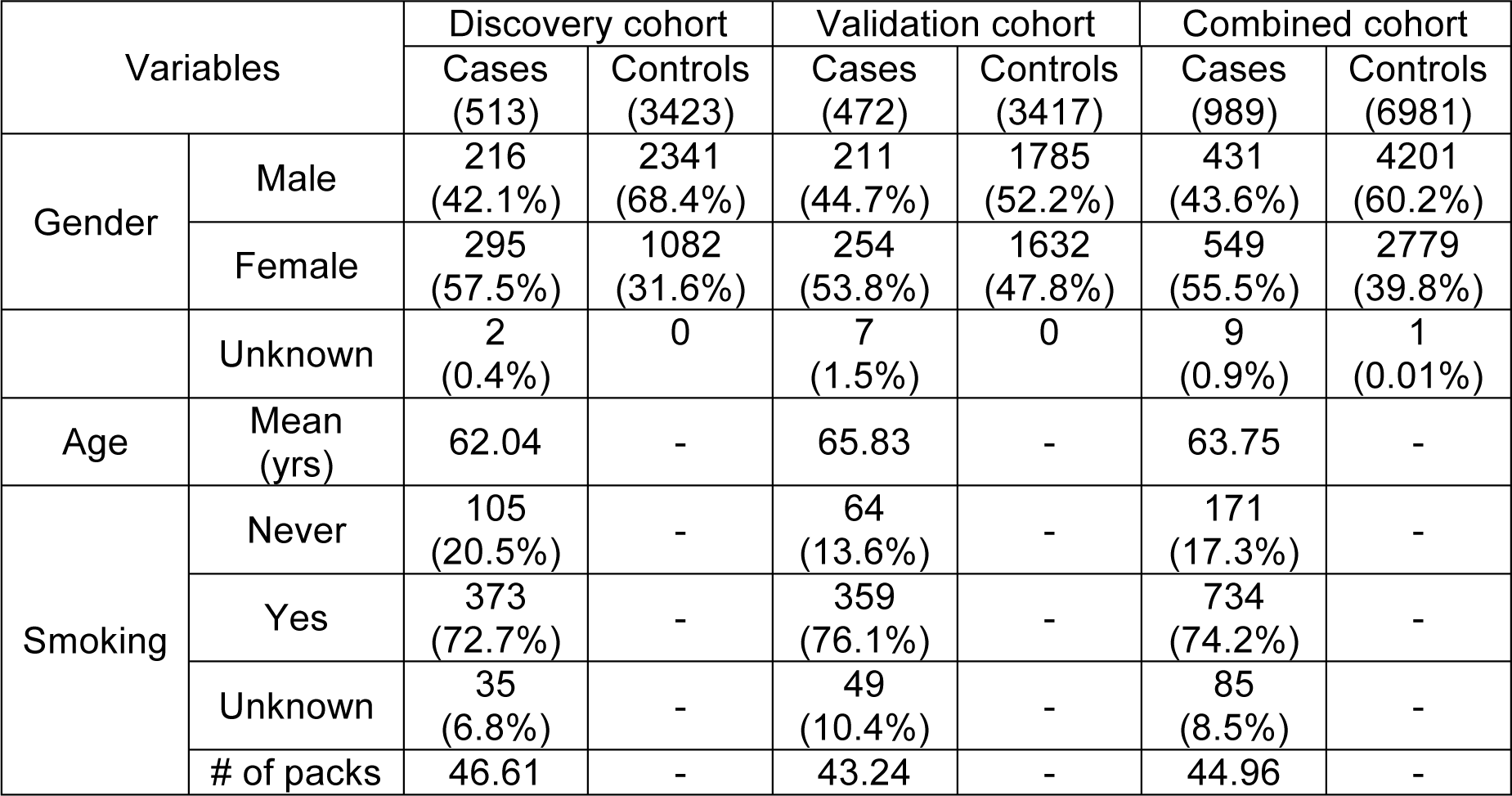
Characteristics of samples in the case-control study cohorts.

### Joint variant calling

We performed variant discovery by realignment and joint variant calling of all case and control germline samples using the GVCF-based best practices for the Genome Analysis Toolkit (GATK, https://www.broadinstitute.org/gatk/) as implemented in a custom pipeline at ISMMS.^12^ Briefly, all samples were independently aligned to human genome build GRCh37 with BWA, subject to indel realignment, duplicate marking, and base quality score recalibration using GATK and Picard, and called to a GVCF file with HaplotypeCaller. Only samples for which over 75% of the exome was callable (depth ≥20, mapping quality ≥10, base quality ≥20) and for which there was no evidence for contamination (VerifyBamID < 3%) were included in the joint variant calling from the GVCF files and variant quality score recalibration with GATK.

## Sample QC

First, we removed samples with >15% of their data missing. We then identified duplicates and related individuals by first or second degree using KING software^13^ and removed a sample from each inferred pair that had the higher fraction of missing data.

Next, we removed any bias that may arise due to systematic ancestry-based variations in allele frequency differences between cases and controls, by adjusting for population stratification using Principal Component Analysis (PCA). Briefly, to identify the population structure, we first removed indels and rare variants (defined by less than 5% of minor allele frequency), using 1000 Genomes^14^ and The Ashkenazi Genome Consortium (TAGC) (https://ashkenazigenome.org) datasets as reference. Then, for the remaining variants, we performed Linkage Disequilibrium (LD) pruning, filtering for a call rate of at least 0.99, and PCA with smartpca using EIGENSOFT 5.0.1 software. Finally, to filter for the least ancestry-based variation in our downstream analyses, we focused on the largest cluster within the PCA plot by PCA gating, which corresponded to individuals of European ancestry. The PCA plots of the discovery, validation and combined case-control cohorts, and the gated regions are shown in Supplemental Figure 1.

### Variant-level QC

For samples that passed the PCA gating, to ensure high-quality genotype/variant calls, we first filtered for variants with: read genotype quality ≥20; read depth ≥10; allelic depth of alternate allele ≥4; variant sites with quality score >50; quality by depth score ≥2; mapping quality ≥40; read position rank sum >–3; mapping quality rank sum >–10 and variant tranche <99%. For heterozygous genotypes, we filtered for alternative allele ratio between 0.30 and 0.70. To reduce differences between case and control samples, we kept sites with differential missingness ≤0.05 between them. Finally, we kept sites with ≥88% of data (in both cases and controls).

### Variant filtering

Next, among the variants that passed QC, we focused on rare, deleterious variants (RDVs) with known pathogenicity. Such variants have been shown to have high penetrance.^15^ To filter out common polymorphisms, we removed any variant present in both case and control cohorts at: minor allele frequencies (MAF) >2%; or in Exome Aggregation Consortium (ExAC) non-TCGA Non-Finnish European population at MAF >1%; or in Genome Aggregation Database (gnomAD) Ashkenazi Jewish population at MAF > 1%. We considered variants that pass these filters to be rare. We filtered the remaining variants for functional impact based on those present in the ClinVar database^16^ using the Annovar tool (http://annovar.openbioinformatics.org). We considered a variant to be pathogenic if it is listed as pathogenic/likely pathogenic in ClinVar; or a frameshift or stopgain variant located 5’ of a variant described to be a pathogenic LOF variant in ClinVar (nonsense and frameshift).

### Statistical analysis

#### Background variation correction

To test for possible background variation between cases and controls, we calculated the tally of rare autosomal synonymous variants per individual. We defined synonymous variants as rare at ExAC MAF ≤0.005% and MAF ≤0.05% in each case-control cohort. Supplemental Figure 2 shows the distribution and background variation statistics of genes with rare synonymous variants in all cohorts. We noted that the frequency of neutral variation varied between cases and controls (Supplemental Figure 2) and accounted for differences in background variation as described below.

**Figure 2:**
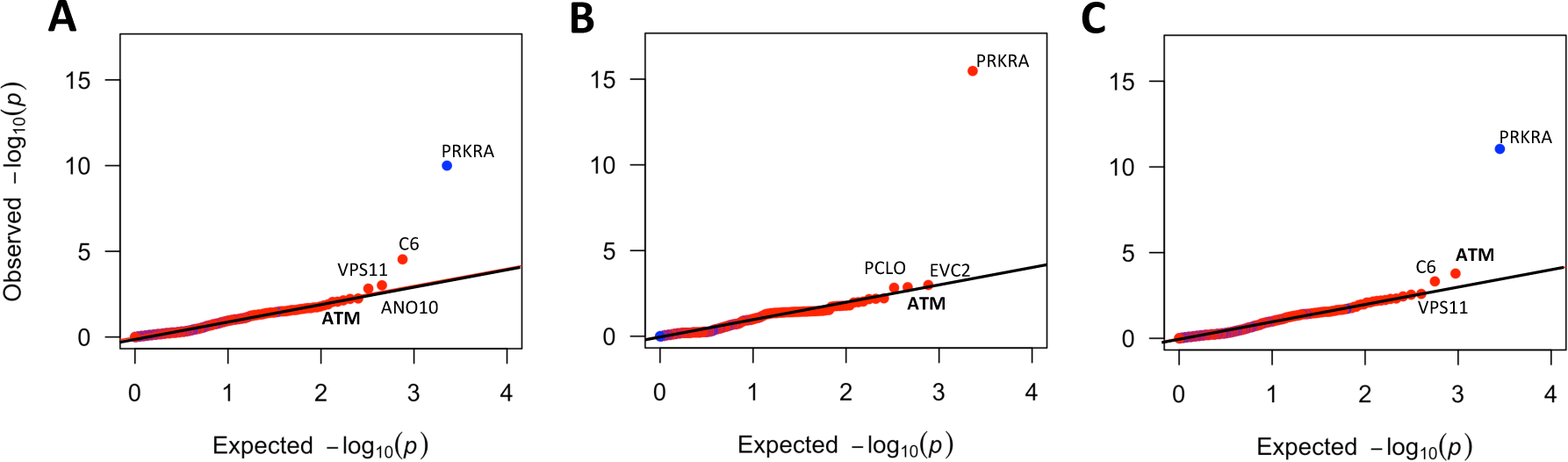
Quantile-Quantile (Q-Q) plots of *RDV* burden test p-values of all genes with RDVs in all study cohorts. **A)** Discovery cohort, **B)** Validation cohort and **C)** Combined cohort. Red represents genes with odds ratio (OR) > 1 and blue represents genes with OR < 1.

#### Gene-level burden analyses

To identify risk genes associated with LUAD, we performed an exome-wide gene-agnostic analysis. First, we filtered for genes above a minimal number of RDVs (cases >2 and controls > 1). In the discovery cohort, 1130 genes had at least one RDV, out of which only 214 passed this filter. In the validation cohort, 258 genes passed this filter. Next, we used aggregate rare, deleterious variant (RDV) burden per gene using Penalized Logistic Regression Analysis (PLRA), within the logistf package in R (https://cran.r-project.org/web/packages/logistf/index.html). To adjust for background variation among samples, we used the number of genes with rare synonymous variants as a covariate for each individual. We deemed genes with *p*-value ≤0.05 and odds ratio >1 as statistically significant risk genes. All statistical tests were two-sided.

#### Enrichment of *ATM* RDVs in a third independent LUAD cohort

Next, to check whether *ATM* is enriched in RDVs in a third independent cohort, we utilized targeted clinical germline sequencing data on *ATM* from 1,594 mostly advanced LUAD cases of European ancestry assayed using MSK-IMPACT (Integrated Mutation Profiling of Actionable Cancer Targets) (Supplemental Method 3).

#### Sex effects on gene-level RDV burden

To test sex effects, we used PLRA with sex as second covariate, and background variation as the first covariate but did not observe statistically significant impact. The percentage of males and females in each cohort are listed in Table 1 (we did not include samples with missing sex data: 9 cases and 1 control).

#### Germline-somatic interactions

To test for interactions of germline variants with somatic mutations, we downloaded somatic mutation data from the Comprehensive TCGA PanCanAtlas,^17^ comprising 465 of the 472 LUAD TCGA cases in the validation cohort. To ascertain mutual exclusivity between *ATM* germline RDVs and somatic *TP53* mutations, we used CoMEt^18^ at default settings.

## RESULTS

### Study Cohorts

To identify genes associated with LUAD risk, we performed an exome-wide multi-stage case-control study, as visually summarized in Figure 1. Briefly, we first performed germline WES on familial-enriched 97 LUAD cases. Then, for increased sample size, we pursued a resource-conscious approach and combined our WES data with those available from cases and healthy controls in dbGaP, for a total discovery cohort of 537 cases and 3,697 controls (see Methods). For validation, we used an independent cohort of 546 sporadic LUAD TCGA cases and 3,953 dbGaP controls (see Methods). We first harmonized the data by realigning and jointly calling germline genetic variants (see Methods). After sample QC, we focused on the largest ancestry-based group for downstream analyses, which were individuals of European ancestry (Supplemental Figure 1). The final discovery dataset included 513 cases and 3,423 controls, while the validation dataset included 472 cases and 3,417 controls. We also considered the combined (discovery + validation) dataset, which after combined QC included 989 cases and 6,981 controls. Clinical characteristics of all cohorts are listed in Table 1.

### *ATM* gene exhibits statistically significant burden of germline RDVs in cases versus controls

Within the filtered cohorts, we focused on *Rare (see Methods) Deleterious Variants (RDVs)*, with deleterious defined as i) being labeled pathogenic or likely pathogenic in ClinVar, or ii) a frameshift or stopgain variant located 5’ of a pathogenic LOF variant in ClinVar (nonsense and frameshift). After QC, we identified at least one RDV in 1,130 genes (median: 1 RDV/gene; range: 1-28) in the discovery cohort. We performed gene-level tests for cumulative RDV burden in cases vs. controls for all genes with RDVs. Figure 2 shows the quantile-quantile (Q-Q) plots of all burden *p*-values. The complete set of genes with burden test *p*<0.05 in the combined cohort is in Supplemental Table 2.

**Table 2:** Gene-level germline rare, deleterious variant (RDV) burden on *ATM* in all study cohorts.

|  | **Discovery cohort** |  | **Validation cohort** |  | **Combined cohort** |  |
| --- | --- | --- | --- | --- | --- | --- |
|  | **Case**<br>**(513)** | **Control**<br>**(3423)** | **Case**<br>**(472)** | **Control**<br>**(3417)** | **Case**<br>**(989)** | **Control**<br>**(6981)** |
|  | ***ATM* Gene** |  |  |  |  |  |
| # Variants | 4 | 7 | 6 | 8 | 9 | 14 |
| # Unique individuals | 5<br>(0.97%) | 8<br>(0.23%) | 7<br>(1.48%) | 9<br>(0.26%) | 12<br>(1.21%) | 17<br>(0.24%) |
| OR (*p*-val)<br>[95% CI] | 4.05 (0.02)<br>[1.3–11.9] |  | 5.50 (1.37e-03)<br>[2.0–14.4] |  | 4.58 (1.66e-04)<br>[2.2–9.5] |  |

From these analyses, we observed that only Ataxia-telangiectasia mutated (*ATM*), a DNA damage repair gene already known to contain moderate-penetrance RDVs predisposing to breast and other cancers,^19,20^ was significantly associated with LUAD in all study cohorts with consistent direction of effect (discovery cohort OR=4.05, *p*=0.02, 95% CI=1.3–11.9; validation cohort OR=5.50, *p=*1.4e-03, 95% CI=2.0–14.4; combined cohort OR=4.58, *p*=1.7e-04, 95% CI=2.2–9.5) (Table 2). We show *ATM* RDVs in Figure 3.

**Figure 3:**
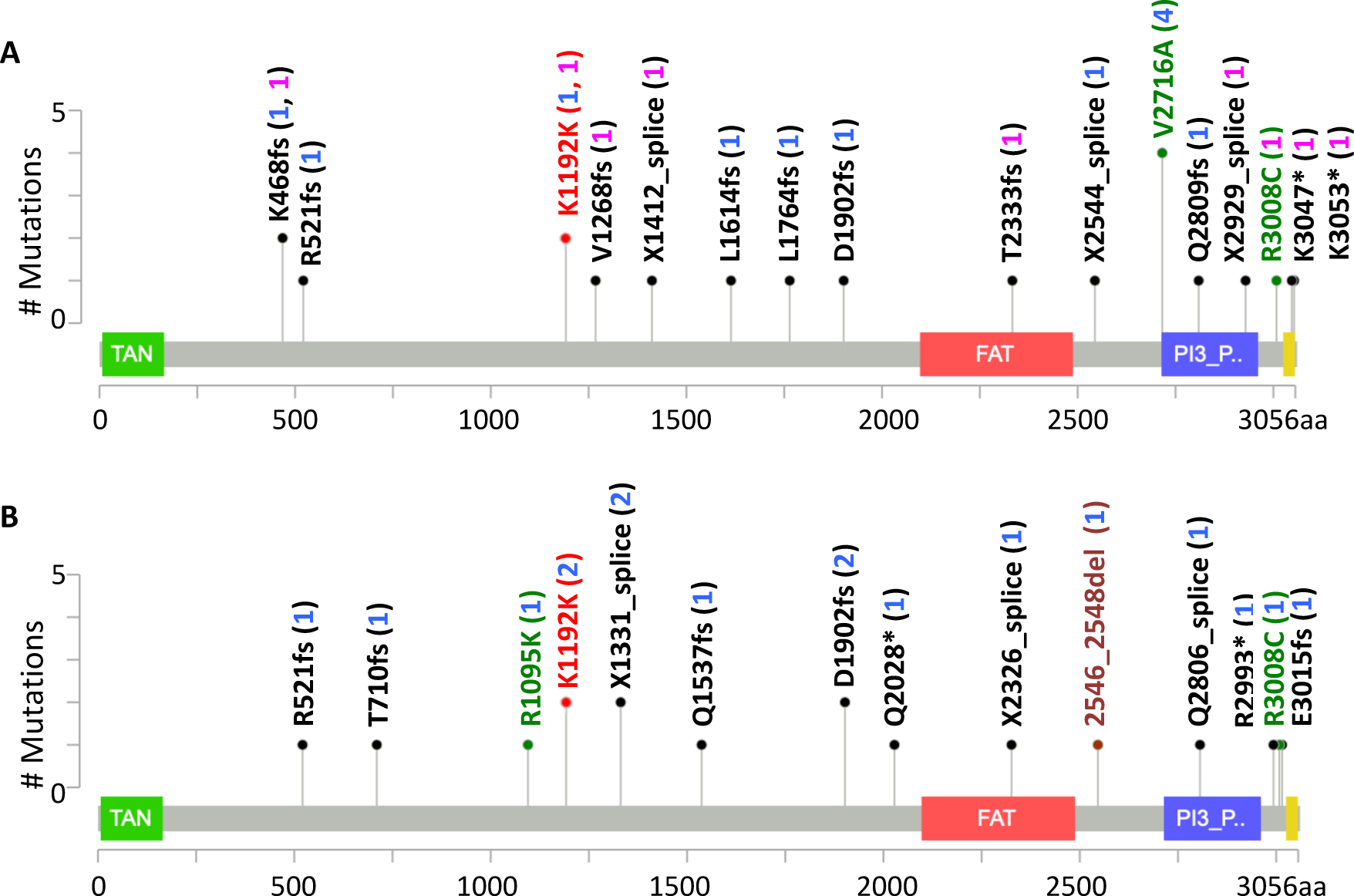
Rare, deleterious ATM variants in all study cohorts. **A)** All cases in the combined cohort plus the MSK-IMPACT cohort (22/2,583) **B)** Controls in the combined cohort (17/6,981). Red: synonymous variants; green: missense variants; maroon: inframe variants; and black: frameshift, splicing and nonsense variants. The variant counts are given in brackets (blue: combined cohort; pink: MSK-IMPACT cohort). 1 intronic variant observed in MSK-IMPACT cohort is not displayed.

Notably, evidencing the importance of using validation cohorts, our rigorous approach enabled us to eliminate genes that were significant in only one cohort or had inconsistent direction of effect between the discovery and validation cohorts. For example, while we observed significant association with risk for *PRKRA* gene in both cohorts, the direction of effect was opposite (discovery cohort OR=0.13; validation cohort OR=3.71; see also Figure 2).

### *ATM* RDVs are enriched in a third independent cohort of 1,**594 LUAD cases**

We next tested enrichment for *ATM* RDVs in an independent set of 1,594 advanced stage LUAD individuals of European ancestry in whom *ATM* was sequenced as part of clinical care at MSKCC (“MSK-IMPACT panel”; see Supplemental Method 3 for clinical details). Consistent with our discovery (0.97% patients; 0.23% controls) and validation cohorts (1.48% patients; 0.26% controls), this clinical cohort also showed enrichment (0.63% patients). Furthermore, we observed that two ultra RDVs from our case cohort were also in the MSK LUAD patients (p.Lys468fs and rs587776551 (p.Lys1192=)) (Supplemental Table 3).

### *ATM* founder variant rs56009889 is more frequent in Ashkenazi Jewish (AJ) cases vs. AJ controls

An *ATM* missense variant, rs56009889 (p.Leu2307Phe), was recently found to be associated with LUAD risk^21^ in individuals of European descent, but we did not include it in our primary analyses due to conflicting pathogenicity information in ClinVar.^16^ While this variant is rare in Europeans, it is relatively common in AJ (gnomAD MAF 3.0%). In our original combined cohort, we found the variant more frequent in cases (MAF 1.06%) than in controls (MAF 0.18%). We then investigated the association between this variant and LUAD in our combined AJ case-control cohort (See Supplemental Figure 3; 120 cases and 284 controls), and observed that it was statistically significantly more frequent in cases (MAF 7.92%) than in controls (MAF 3.17%) (OR=2.65, p=0.007, 95% CI=1.3– 5.3) (Supplemental Table 4).

### *ATM* germline RDV carriers with Loss of Heterozygosity (LOH)

Several recent studies suggest that in breast,^19,22^ pancreatic^23^ and prostate cancer patients with heterozygous germline pathogenic *ATM* variants, the remaining wild-type copy is frequently inactivated (40% to 79%). To determine whether LOH was also common in LUAD patients with *ATM* RDVs, we investigated the 7 individuals with *ATM* RDVs for whom tumor data was also available (3 males and 4 females in TCGA). We observed the same trend, with 3/7 (43%) patients exhibiting LOH (rs587779846 (p.Leu1764fs); rs587779866 (c.7630-2A>C, splice); and rs587782652 (p.Val2716Ala)). None had second somatic hits at other *ATM* coding loci. Of the four non-LOH patients, two had somatic mutations in the ATM-interacting protein, EphA5.

### *ATM* germline RDV carriers have distinct somatic mutational patterns that are mutually exclusive of *TP53* mutations

We next asked whether similar to recent studies on *ATM* germline variant carriers with breast cancer,^22^ the 7 patients with *ATM* germline RDVs showed differences in their somatic mutation patterns compared to 465 non-carriers in TCGA data. In carriers, top recurrent somatically mutated genes were *LRP1B* (71.4%), *KRAS* (57.1%), *EPHA5* (28.6%), *PTPRS* (28.6%), *STK11* (28.6%) and *TRRAP* (28.6%). However, for non-carriers, the top somatically mutated gene was *TP53* (52.2%), followed by *LRP1B* (33.8%) and *KRAS* (29.5%). Notably, consistent with observations in breast cancer, only one *ATM* RDV carrier had a *TP53* somatic mutation (14.3%); in fact carrier status was mutually exclusive of somatic *TP53* mutations (p=0.03, Supplemental Figure 4). While our study population is relatively small, consistent with other studies,^20,22^ it suggests germline *ATM* RDVs impact somatic mutational patterns in LUAD individuals, which could have clinical implications.

## DISCUSSION

To identify genes associated with increased risk for LUAD, we have performed by far the largest population-based study on germline WES datasets to date, reporting results on 1,083 cases and 7,650 controls. Our approach has several unique advantages. First, we explored the genetic basis for LUAD predisposition in an unbiased exome-wide manner rather than performing a candidate-gene based approach that only investigates a few genes.^24^ Second, using WES datasets enabled us to investigate *rare* variants, which can have higher penetrance than *common* variants typically discovered in GWAS studies^15,25^ which we filtered for using a strict pathogenicity criteria based on ClinVar.^16^ Third, unlike prior WES studies, we jointly analyzed case and control WES together, which enabled us to avoid biases associated with using population-level databases as controls (e.g. such databases do not allow us to correct for individuals with multiple rare variants). Finally, this is the first LUAD WES study that validates results in an independent case-control validation cohort, which we even checked in a third independent cohort within a clinical setting. Our results rigorously establish that, in addition to its known role as a predisposition gene for other cancers including pancreatic ductal adenocarcinoma,^26^ breast^27^ and prostate,^28^ ATM is a LUAD predisposition gene.^29,30^

As we have used the TCGA LUAD cases in our validation cohort, it is worth mentioning that this cohort has been analyzed in recent other studies.^20,30–32^ However, these studies were limited in scope. Parry *et al*^32^ studied 8 DNA repair genes and observed that *ATM* had the highest number of rare pathogenic germline variants, while Lu *et al*^20^ observed that *ATM* had the highest number of rare *truncating* germline mutations, both only studying TCGA LUAD cases. A recent study^30^ suggested that *ATM* RDVs were enriched in LUAD cases,^33^ but only compared case variant frequencies to population database controls, without validation in any independent case-control cohort. These findings are further complemented by a study on *ATM common* variants in a case-control cohort,^34^ which identified significant association with lung cancer risk in never-smokers.

One germline *ATM* variant, rs56009889, that others recently associated with LUAD risk^21^ in individuals of European ancestry, shows conflicting interpretation of its pathogenicity in ClinVar.^16^ Therefore, we did not include it in our primary analysis. To study this variant further, which is a founder variant in the AJ population, we focused on the AJ individuals in our combined cohort. Our results support the previous reports on this variant as a risk variant. Given its high frequency in a particular population, further studies are needed to assess its pathogenicity and evaluate its importance for inclusion in risk assessment clinical genetics testing.

This study should be interpreted in the context of potential limitations. We were unable to perform risk stratification based on smoking history due to incomplete smoking information for most controls. As ∼70% of individuals with LUAD smoke, well-annotated control samples will enable a better understanding of the effects of smoking and other environmental agents as confounders on germline risk for these distinct lung cancer subtypes. We anticipate that as WES databases such as dbGaP gets populated, and more data get published in large studies such as UK Biobank, future studies will address such limitations.

To conclude, while lung cancer has a dismal survival rate, it can be prevented, managed or treated by the timely detection of individuals at high-risk. Here, we used population-based sampling of case-control individuals of European ancestry to identify genetic markers of LUAD risk and demonstrated that individuals with *ATM* germline RDVs are at increased risk. As *ATM* is also a recognized risk gene for cancers of the pancreas, breast and prostate, this finding suggests that LUAD may be a part of the *ATM*-related cancer syndrome. Furthermore, as individuals with *ATM* variants have increased surveillance for these cancers to increase early inception, our findings have important implications for their additional surveillance for LUAD with low-dose CT (as is done for individuals with a history of heavy smoking).

Notably, LUAD individuals with *somatic ATM* variants have favorable treatment outcomes for local response to radiotherapy (RT)^35^ and immunotherapy.^36^ These strongly support future research efforts towards understanding the association of *ATM* germline RDVs with treatment outcomes, which would strongly impact the cost/benefit analyses for clinical genetics testing.

## Data Availability

We downloaded the TCGA germline WES BAM files from National Cancer Institute Genomic Data Commons (GDC) data portal and the WES files for other samples from dbGaP. We will deposit the WES data for 97 samples that we sequenced in dbGaP.

## ACKNOWLEDGEMENTS

This work was supported by a grant to Z.H.G from LUNGevity Foundation and in part through the computational resources and staff expertise provided by Scientific Computing at the Icahn School of Medicine at Mount Sinai. We thank Dr. Charles Powell for help with MSSM IRB protocol.

## AUTHOR CONTRIBUTIONS

M.E., R.J.K. and Z.H.G. wrote the manuscript. M.Z., C.R., S.M.L. and G.R. recruited patients. V.V. and S.J. led sample sequencing at PGDx. R.J.K. and Z.H.G. performed germline sequence analysis (variant calling). M.E. performed burden and other data analyses. S.M and K.Of. analyzed MSK-IMPACT cohort data. K.On. interpreted results.

Z.H.G. conceived, designed and supervised the study. All authors approved the final manuscript.

## Notes

**FUNDING:** This work was supported by a grant from LUNGevity Foundation.

### Competing Interest Statement

The authors have declared no competing interest.

### Funding Statement

This work was supported by a grant from LUNGevity Foundation.

